# Estimating E-Cigarette Use Prevalence Among US Adolescents Using Vaping-Related Online Search Trends

**DOI:** 10.1101/2020.11.21.20236133

**Authors:** Floe Foxon

## Abstract

**Introduction:** Adolescent e-cigarette use is a developing phenomenon. Greater surveillance of underage use is necessary to inform e-cigarette policy and mitigate adolescent e-cigarette use. Accurate prevalence estimates for adolescent e-cigarette use are provided by large national surveys. However, these surveys are costly and provide only annual estimates. To obtain more affordable estimates faster and more frequently, novel methods are required.

**Methods:** Online search term popularity data were taken from Google Trends. Interest in the search terms ‘vapes’, ‘vape’, and ‘vape pen’ were followed monthly from January 2011 to November 2020. Time-lagged zero-normalized cross-correlations were performed between the Google data and current (past 30 day) high-school e-cigarette use prevalence estimates from the National Youth Tobacco Survey (NYTS). The search interest data were then calibrated to the NYTS data to estimate adolescent e-cigarette use prevalence using online searches.

**Results:** Maximum correlation coefficients of 0.979 for ‘vapes’ and 0.938 for ‘vape’ were obtained when search interest lagged use prevalence by one month, and 0.970 for ‘vape pen’ when the lag was two months (*p <* 0.0001 for all). Calibrating the search term data to NYTS provided a high-school current e-cigarette use prevalence estimate of 11.3–16.5% for November 2020, suggesting adolescent use of e-cigarettes has continued to decline since the NYTS estimate of 19.6% for January–March 2020.

**Conclusion:** Online search trend data may provide reasonably reliable and more frequent estimates of adolescent e-cigarette use prevalence at substantially lower costs than traditional surveys. Such additional data may help in developing adolescent e-cigarette use prevention efforts.

## Introduction

E-cigarettes have emerged as an alternative nicotine source and possible future smoking-cessation device for adult smokers (American College of Cardiology, 2020; Hajek et al., 2019), but adolescent use of e-cigarettes (Gentzke et al., 2019) presents a public health challenge.

Tobacco control policies are shaped by available data on prevalence of product use. Existing data sources for adolescent e-cigarette use prevalence include the Youth Risk Behavior Survey (YRBS) (US Centers for Disease Control and Prevention, 2020) and the National Youth Tobacco Survey (NYTS) (US Centers for Disease Control and Prevention, 2019a) both administered by the US Centers for Disease Control and Prevention, as well as various state-wide surveys such as the state Youth Tobacco Surveys (YTS) (US Centers for Disease Control and Prevention, 2019b) and Healthy Kids surveys (California Department of Education, 2020; Colorado Department of Public Health & Environment, 2019).

These surveys utilize large sample sizes and complex sample designs which provide the best existing estimates of adolescent e-cigarette use prevalence. However, administering state- and nation-wide surveys is a costly endeavor in terms of the time, money, and labor involved. Consequently, these surveys are administered at most annually or biannually.

Because adolescent e-cigarette use is evolving, data with higher temporal resolution (i.e. less time between point estimates) and which require fewer resources to collect are necessary; waiting an entire year to determine the impact of a policy may risk irreversible consequences if that policy fails or backfires.

Adolescent e-cigarette use intersects with the online world. Half of US adolescents may be exposed to tobacco or e-cigarette related social media (Hebert et al., 2017), and about 1 in 10 adolescent e-cigarette users source the e-cigarettes they use from the internet (Merianos et al., 2019). E-cigarette related discussion forums have also been identified as a rich source for information on adolescent e-cigarette use behavior (Zhan et al., 2019). Given then that adolescent e-cigarette use is associated with online content, this raises the question of whether vaping-related internet use predicts real-world e-cigarette use.

Google Trends (Google, 2020), an online, search-term-popularity data generator, has been demonstrated to have modest reliability in reflecting geographical and temporal patterns in epidemiological settings (Cervellin et al., 2017). Extant research has applied Google Trends data to predict outbreaks of influenza (Carneiro & Mylonakis, 2009) and COVID-19 (Ahmad et al., 2020). In nicotine and tobacco science, relative Google search volumes have been compared for heat-not-burn products and e-cigarettes to investigate heat-not-burn market growth (Caputi et al., 2017). Most relevant to the present study, statistically significant but weak-to-moderate strength geographical correlation was demonstrated between Google Trend data and state use prevalence for cigars (Cavazos-Rehg et al., 2015). This study aims to determine whether Google Trends data correlates temporally with adolescent e-cigarette use prevalence in the US using NYTS prevalence estimates from 2011–2020. This presents novel research which may provide faster and higher temporal resolution estimates of adolescent e-cigarette use prevalence.

## Methods

### Sample

National estimates for the prevalence of past 30 day (current) e-cigarette use among US high-schoolers (here ‘adolescents’) from 2011–2020 were taken from resources published by the US Department of Health and Human Services (US Food and Drug Administration, 2019b; Wang et al., 2020). These prevalence estimates are derived from NYTS, which is an annual, nationally representative survey of US adolescents with a multi-stage probabilistic cluster sample design. Approximately 17,000–25,000 adolescents were surveyed at each wave. The exact dates through which these surveys were administered were taken from their annual methodology reports (US Centers for Disease Control and Prevention, 2019a).

Online search popularity data were taken from Google Trends. Google Trends provides normalized ‘search interest’ values which represent the popularity of a given search term through Google web searches on a scale from zero to 100. A value of 100 represents the peak popularity for that term, which falls on some month. A value of 50 on another month means the search term was half as popular on that month than it was in the month when the value was 100. Monthly data for the vaping-related search terms ‘vapes’, ‘vape’, and ‘vape pen’ were collected from January 2011 to November 2020. The search terms ‘vapes’ and ‘vape pen’ were selected because they are listed as alternative names for e-cigarettes in the 2019 NYTS questionnaire definition (which precedes the e-cigarette-related questions in this survey) (US Centers for Disease Control and Prevention, 2019a).

The range of dates was selected to coincide with NYTS data collection, and the data were restricted geographically to ensure only searches from within the United States were included for analysis.

### Analyses

The NYTS prevalence estimates were normalized, and the Google web search interest data were re-normalized by dividing each element in each set of data by the maximum value in that set of data such that for all three sets, a value of one represented the maximum value in each set. These data were then plotted to observe the similarities and differences in their trends over time.

The midpoint dates between the beginning and end of survey administration for each NYTS wave were found, and the months these dates fell on were taken to be the months which correspond to the prevalence estimate for that year. The NYTS data were then paired with the Google web search data for the same months (zero lag). Treating the data as signals over time, zero-normalized cross-correlations were performed between NYTS and each of the Google web searches with these month-pairs. This was then repeated with month time lags up to three months in either direction, for example a one month lag was implemented by cross-correlating the NYTS data with Google Trends data which correspond to the NYTS months minus one month, and a ‘-1’ month lag was implemented by cross-correlating the NYTS data with Google Trends data which correspond to the NYTS months plus one month. Corresponding *p*-values were calculated for the cross-correlation coefficients with statistical significance at *alpha* = 0.05.

Finally, the peaks of the search term popularity trends were calibrated to the peak NYTS e-cigarette use prevalence (27.5%) such that e-cigarette use prevalence estimates could be obtained from the search term data for any given month. In this way, prevalence estimates were obtained for November 2020.

All analyses were conducted in Python version 3.7.6 with the packages NumPy version 1.18.5, Matplotlib version 3.2.2, and Pandas version 1.0.5. Code will be made available upon reasonable request.

## Results

Figure 1 shows the normalized trends in high school e-cigarette use prevalence and Google web search popularity for the search terms ‘vapes’, ‘vape’, and ‘vape pen’ over time. It can be readily seen than all three trends display very similar shapes.

**Figure 1:**
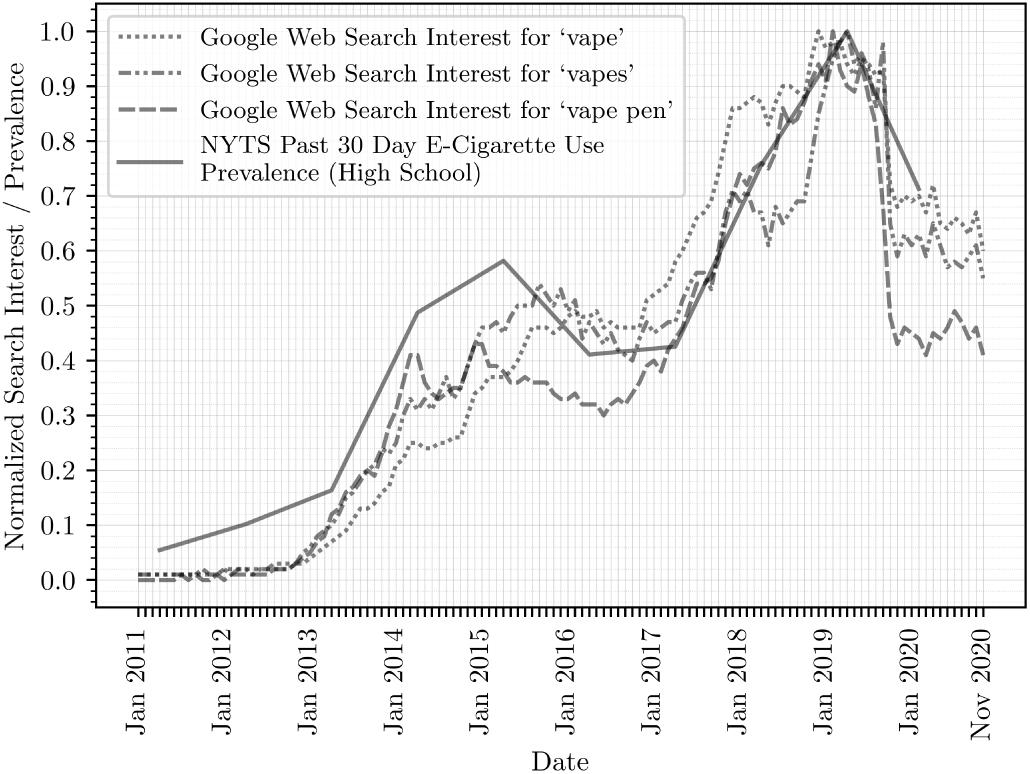
Trends in Vaping-Related Online Searches and Survey-Measured Prevalence of High School E-Cigarette Use. The dotted and dashed lines show the change in normalized search interest over time for the vaping-related Google web search terms ‘vape’, ‘vapes’, and ‘vape pen’. A normalized search interest value of 1.0 represents the peak popularity for that term. A value of 0.5 means the search term was half as popular than it was at 1.0. The solid line shows the normalized prevalence of current (past 30 day) e-cigarette use among US high school students from NYTS. A value of 1.0 means the peak prevalence of current e-cigarette use among this cohort (27.5% in 2019). A value of 0.5 means current e-cigarette use was half as prevalent as at 1.0.

Table 1 shows the zero-normalized cross-correlations between the NYTS prevalence estimates and corresponding Google web search interest for a range of time lags. The maximum cross-correlation coefficients were 0.979 for ‘vapes’ and 0.938 for ‘vape’ with search interest lagging NYTS prevalence by one month, and 0.970 for ‘vape pen’ with search interest lagging NYTS prevalence by two months. Across the entire range of lags, all cross-correlations for all terms were statistically significant.

**Table 1:** Time-Lagged Zero-Normalized Cross-Correlations Between NYTS Prevalence and Google Search Interest.

| Number of Months<br>Search Interest Lags<br>NYTS Prevalence By | Cross-Correlation Coefficients |  |  |
| --- | --- | --- | --- |
|  | ‘vapes’<br>Search<br>Term | ‘vape’<br>Search<br>Term | ‘vape pen’<br>Search<br>Term |
| 3 | 0.960** | 0.931** | 0.966** |
| 2 | 0.967** | 0.934** | **0.970\*\*** |
| 1 | **0.979\*\*** | **0.938\*\*** | 0.966** |
| 0 (no lag) | 0.972** | 0.929* | 0.958** |
| -1 | 0.967** | 0.923* | 0.945** |
| -2 | 0.969** | 0.925* | 0.940** |
| -3 | 0.963** | 0.916* | 0.915* |

Bold font indicates maximum correlation
\*\* $p < 0.0001$ \* $p < 0.001$

Calibrating the search term popularity to the NYTS prevalence data gave current e-cigarette use prevalence estimates of 15.1% (‘vapes’ search term), 11.3% (‘vape pen’ search term), and 16.5% (‘vape’ search term) for November 2020.

## Discussion

This research aimed to determine whether online vaping-related search data from Google Trends correlates with adolescent e-cigarette use prevalence in the US. The results presented here show very high and statistically significant correlation between NYTS prevalence estimates and interest in the search terms ‘vapes’, ‘vape’, and ‘vape pen’.

Correlations were strongest with 1–2 month lags, suggesting changes in vaping-related search trends are shortly followed by changes in adolescent e-cigarette use trends, however whether this relationship is causal is not determined.

Using the search trend data to predict high school current e-cigarette use prevalence provided a prevalence estimate for November 2020 of 11.3–16.5%, which is lower than the 19.6% estimated by NYTS for January–March 2020. These findings may show that the number of adolescent e-cigarette users has continued to decline since NYTS 2020 was administered. This suggests that recent adolescent e-cigarette use prevention efforts such as Tobacco 21 (US Food and Drug Administration, 2019a), along with likely effects from pandemic-mitigation efforts, have had some impact on reducing adolescent e-cigarette use.

Although a proof-of-concept, this research shows that monitoring online interest in vaping may prove to be a cost-free and reliable means of generating more frequent prevalence estimates than large national surveys.

Future research may apply these methods to specific geographies and patterns of use.

### Strengths

Strengths of this research include the novel application of temporal online search data to predict trends in e-cigarette use prevalence among US adolescents, and rigorous statistical analysis.

### Limitations

The primary limitation of this research is that the mechanism which associates vaping search interest with e-cigarette use prevalence is complex, therefore an increase in search popularity does not necessitate an increase in use prevalence. Online dissemination of information regarding the health risks of nicotine product use may result in greater search popularity but not greater use prevalence. Never e-cigarette users may search vaping-related terms out of curiosity without initiating e-cigarette use, and former users may search for support to sustain their abstinence. Furthermore, Google Trends does not provide age-filtering of

Google web searchers, therefore adolescent vaping-related search trends are confounded by other age groups making the same searches. However, US internet usage is disproportionately dominated by younger Americans (Jones Fox, 2009).

## Data Availability

NYTS data are publicly available from the US Centers for Disease Control and Prevention. Google Trends data are publicly available from Google.

https://www.cdc.gov/tobacco/data_statistics/surveys/nyts/data/index.html

https://trends.google.com/trends/?geo=US

## Acknowledgements

The author would like to acknowledge Dr Arielle Selya for proof-reading this article, and Joe Gitchell for providing insightful comments and suggestions.

